# Essential information about chronobiology and chronotherapy for the optimal care of people with bipolar disorders: an international expert consensus

**DOI:** 10.1101/2025.06.17.25329750

**Authors:** Jacob J. Crouse, Victoria Loblay, Anthony Jorm, Timothy Wong, Zsofi de Haan, Carla Gorban, Mirim Shin, Emiliana Tonini, Chris Aiken, Lauren B Alloy, Kürşat Altinbaş, Serge Beaulieu, Joanne S Carpenter, Bruno Etain, Yuichi Esaki, Jess G. Fiedorowicz, Corrado Garbazza, Pierre A Geoffroy, Bartholomeus C.M. Haarman, Tone E.G. Henriksen, Maria Paz Hidalgo, Maree L. Inder, Raymond W. Lam, Helle Ø Madsen, Colleen A. McClung, Gunnar Morken, Laura Palagini, James Phelps, Richard J. Porter, Aswin Ratheesh, Rixt F. Riemersma-Van der Lek, Janusz K. Rybakowski, Erika FH Saunders, Peter F.J. Schulte, Daniel J. Smith, Holly A. Swartz, Bryan K. Tolliver, Anna Wirz-Justice, Ian B Hickie, John F Gottlieb, the Chronobiology Task Force of the International Society for Bipolar Disorders

## Abstract

**Background:** Circadian dysfunction is involved in the pathophysiology of bipolar disorders (BD), and circadian-based interventions are gaining recognition in their management. Moreover, basic and epidemiologic research has generated findings inspiring circadian-informed self- and clinician-management strategies. Despite these gains, many Clinical Practice Guidelines and clinical training programs have not incorporated this evidence in their recommendations and curricula.

**Aims:** This International Society for Bipolar Disorders (ISBD) Chronobiology and Chronotherapy Task Force position paper reports a Delphi study-based international consensus statement on what is essential for mental health clinicians to know about the chronobiology and chronotherapy of BD.

**Methods:** An initial pool of statements was derived via review of academic and grey literatures. Statements were rated on a 5-point scale (essential; important; don’t know/depends; unimportant; should not be included). Consensus was reached when statements were rated as essential or important by ≥80% of experts. Two young persons with lived experience of BD gave feedback on acceptability.

**Results:** Thirty experts from 15 countries in Europe, North and South America, and the Asia Pacific participated (mean age 55.3 years [SD=11.8]; 40% female; 83% psychiatrists; mean clinical and research experience of 26 [SD=10.8] and 22 years [SD=12.6], respectively). Eight-hundred-and-forty-seven ratings occurred across 12 core constructs and three rounds; 470 statements were discarded after one round and 35 were discarded after rerating. Consensus was reached on 342 statements spanning four themes: basic circadian science; circadian health and disruption; chronobiology of BD; and six chronotherapies (e.g., key protocols, outcomes, side effects, risks/contraindications).

**Conclusions:** This position paper summarises an international consensus statement on the essential information about the chronobiology and chronotherapy of BD that is intended to help clinicians optimise their management of individuals with BD. This knowledge is available online (https://osf.io/agk9y) and its dissemination is expected to enhance the training and efficacy of clinicians globally.

## Background

There are several models of the pathophysiology of bipolar disorders (BD) that inform diagnosis and treatment. These include disturbances in neurotrophic and neuroinflammatory processes, mitochondrial bioenergetics, monoamine signalling, and reward function^1–3^. Over the past 50 years, the basic science of chronobiology and the applied field of chronotherapeutics has generated another foundational model for understanding and treating BD^1,2,4,5^. In the 1970s-1980s, evidence emerged suggesting that disturbances in circadian rhythms contributes to the pathophysiology of BD^6–10^. Supportive evidence has since accumulated across many approaches: genetic^4^, family^11,12^, epidemiologic^13^, longitudinal^14,15^, physiologic^4,16–19^, biobehavioural^4,14,20–22^, and therapeutic^23^. The hypothesis of a core role of circadian rhythms in BD is now more widely accepted^4^.

### Gaps in guidelines

Despite these gains, dissemination of information to clinicians about the chronobiology and chronotherapeutics of BD remains limited. This gap is clearest for Clinical Practice Guidelines (CPGs). A review of 25 CPGs for BD found that fewer than half included even a basic discussion of circadian disturbances^24^. The authors concluded: “*Given the evidence regarding the importance of circadian dysrhythmias in the onset, course and prognosis of BD, we suggest that omission of information and practical advice on management needs addressing in future updates of CPGs for BD*” (p. 444)^24^. This review highlighted a contrast between promising data about chronotherapeutics for BD^23^ and limited discussion in CPGs of circadian dysfunction as a pathophysiology and treatment target. For example, in the case of Bright Light Therapy there are at least seven randomised controlled trials (RCTs) suggesting its efficacy, safety, and tolerability in the treatment of bipolar depression^25^, and there is RCT evidence for Interpersonal and Social Rhythm Therapy^26,27^ and dark therapy using blue-blocking glasses^28^. Some CPGs give attention to chronobiology regarding the etiology and pathophysiology of BD (e.g., 2020 Royal Australian and New Zealand College of Psychiatrists CPG^29^) whereas others do not; for example, the words “circadian”, “chronobiology”, and “chronotherapy” do not appear in the 2018 CANMAT/ISBD CPG^30^ (although Bright Light Therapy is discussed regarding bipolar depression).

In addition to RCT evidence, a wealth of clinically informative findings comes from observational studies which encourage circadian-based, self-management strategies (e.g., behavioural regulation of the timing of sleep, wake, and light exposure). Observations from naturalistic and cohort studies that are missing from, or minimised in, CPGs may help clinicians manage and anticipate their patients’ course of illness. For instance, knowing a patient’s chronotype can inform prognosis and treatment: higher morningness chronotypes tend to respond better to lithium, whereas higher eveningness chronotypes have lower success with such treatments, and are more likely to present with features like suicidality^31^. Knowing that dampened circadian rhythms are often a core feature may prompt inclusion of rhythm-boosting strategies into treatment plans. And identifying a seasonal pattern of illness can help clinicians plan the timing and intensity of strategies and treatments during destabilising periods.

### Gaps in training

Although suggested only by sparse data, we believe that chronobiologic information is neglected in clinical training programs. In the Box, a person with lived experience of BD, who was involved in this study, recounts their story of BD and the lack of exposure to information about chronobiology/chronotherapy in their encounters with Australian medical practitioners. Of the published evidence, a survey of 409 medical schools in 12 countries reported that the average amount of time spent on sleep education is <3-hours, with around one-quarter of respondents suggesting an absence of sleep curricula^32^. A survey of Brazilian psychologists reported that 76% had no academic exposure to biological rhythms, 67% were unfamiliar with the term ‘chronobiology’ (introduced 50+ years ago), and 63% could not describe a biological rhythm beyond the sleep-wake cycle^33^. A review suggests that a neglect of sleep in training programs is widespread across disciplines^34^. Relatedly, the Society for Research on Biological Rhythms recently endorsed the need for a *Circadian Medicine Course* across health and medical fields^35^; there may be value in BD-specific curricula.

### Current Study

The science of chronobiology and chronotherapeutics in BD is complex. There is no consensus to guide what information should be disseminated to help clinicians optimise their care of people with BD. While clinician’s guides (e.g., ^36,37^) and many online articles (of variable quality) summarise strategies to manage sleep-wake cycles and circadian rhythms in BD, the amount of information in this area is vast^4^. Accordingly, this *International Society for Bipolar Disorders* (ISBD) *Chronobiology and Chronotherapy Task Force* position paper reports an expert consensus (created via Delphi methodology) on the *essential* information for clinicians supporting people with BD to know about the chronobiology and chronotherapy of BD to optimise their care.

**Box: Perspective by a young person with lived experience of bipolar disorder*.**

I would count myself one of the lucky ones. As soon as I was diagnosed with bipolar disorder, I received the best available care. This was because it was informed by the evidence that circadian rhythms play a key role in bipolar, which is essential to understand if you are going to treat the condition effectively. From my treating specialist, I learnt how I could drive my own care by engaging in circadian-based interventions such as waking up at the same time every morning and taking walks in the morning sunlight. I came to understand how these self-initiated strategies could regulate my mood and improve my sleep. Knowing that bipolar symptoms can be exacerbated by seasonal changes in light, my specialist and I can closely monitor my symptoms during these periods and draw on chronotherapy, not just as a treatment for symptoms, but as a protective intervention. Understanding that bipolar is linked to physiological changes in circadian rhythms within my body, I can also comprehend that my mental health condition is not who I am, but a physical illness that can benefit from medical treatment and self-management, no different from any other complex physical illness, such as diabetes or hypertension.

Sadly, while my first symptoms of bipolar disorder occurred when I was just 14 years old, I did not receive the accurate diagnosis of bipolar disorder, and access to this appropriate chronobiology-informed treatment until I was 25. This caused great devastation in my life, and unnecessarily prolonged my suffering. I have often wondered whether this subsequent delay in appropriate treatment may have been avoided had the health professionals I interacted with along my journey understood the role of circadian rhythms in bipolar and how this presents.

It may come as a surprise that I was not diagnosed until I was 25 years old, given that I saw my first psychiatrist when I was just 14 and continued to engage with psychiatrists regularly from that point in time. I presented as a young adolescent to both general practitioners and psychiatrists with what I now know to be classic circadian depression symptoms associated with bipolar, including crippling fatigue, impaired sleep quality and concentration, and psychomotor retardation, among others. If these symptoms had been assessed through a chronobiology lens, I find it difficult to believe that I would not have received a proper diagnosis and optimised treatment at an earlier stage in my life. This may have spared me and my loved ones a great deal of pain, allowed me to learn how to self-manage my condition through psychoeducation of circadian-based strategies, and reduced self-stigma and the stigma of others.

The health professionals I have interacted with across my life have been well-intentioned but provided under-informed care. While they have sought to offer compassionate care it has been hindered by profound knowledge gaps comprised of a lack of understanding of the role of chronobiology in bipolar disorder. I believe an attuned health professional with knowledge of the biology of bipolar would have been able to easily assess and identify my condition a mile away, provided appropriate chronotherapeutic interventions, and empowered me to drive my own care. Had this taken place when I first started experiencing my symptoms this would have been life changing, in the same way I have flourished since my care began being informed by chronobiology in my mid-twenties. For the first time since I got sick, I feel like me again.

*Note: This person has clinical and research training and works for/with the team in a salaried capacity.

## METHODS

### Ethical approval

The study had ethical approval from the University of Sydney Human Research Ethics Committee (2023/981). Prospective participants were given a Participant Information Sheet and told that advancing through the questionnaire implied consent to participate.

### Study design

The study followed the key elements of the Delphi method^38^. First, the facilitators designed and organised the Delphi study. They compiled a questionnaire with a list of statements about BD and chronobiology or chronotherapy for experts to rate for agreement. A statistical criterion was chosen to define consensus among the aggregated expert ratings. Experts could suggest new statements in the initial round for rating in subsequent rounds. The facilitators shared anonymous feedback to the experts about how their ratings compared to the rest of the panel. Experts could revise their ratings after receiving this feedback. Ratings were aggregated across rounds, with the statistical criterion being used to define consensus and to determine which statements needed re-rating in a later round.

### Creation of the initial statements (Round 1)

The facilitators extracted statements for Round 1 of the study from three sources: (1) reviews published by the ISBD *Chronobiology and Chronotherapy Task Force*^4,23,39^; (2) a systematic search of primary studies, systematic reviews, and meta-analyses on BD and chronobiology or chronotherapy published after 2019 (after the first ISBD *Chronobiology and Chronotherapy Task Force* review); and (3) a systematic search of grey literature on the topic (see Appendix, p. 2). There was no *a priori* limit on the number of statements.

Nine-hundred-and-sixty statements were extracted and refined by rephrasing for clarity, synthesising conceptually similar statements, or deleting those out of scope. This resulted in 758 statements for rating in Round 1.

### Recruitment of experts

The facilitators contacted representatives from ISBD, Society for Light Treatment and Biological Rhythms, and the Center for Environmental Therapeutics, who sent an email invitation to their memberships. Potential experts were guaranteed co-authorship on the study publication if they completed all rounds (maximum=3) of the Delphi study. Recipients of the invitation were asked to send it to relevant colleagues.

There is no formal sample size calculation for Delphi studies^40^. Publications suggest that the results of single stakeholder Delphi studies stabilise^38^, and achieve moderate replicability, with 20-30 participants^40^; accordingly, a required sample size of 30 experts was set for Round 1.

### Questionnaire

The questionnaire was deployed using REDCap. The landing page included the Participant Information Sheet and an eligibility screener, with inclusion criteria being: (i) aged ≥18; and (ii) clinician with ≥10 years treating BD; or (iii) researcher with ≥3 first author papers or ≥5 total papers in the area (BD and chronobiology/chronotherapy).

Experts then presented with instructions on how to rate the statements. They were told that their responses were anonymous and were asked, for each statement, to consider the question: “*How important is it that a clinician treating people with bipolar disorder knows this information*?” Statements were rated on a 5-item Likert scale: “Essential”, “Important”, “Don’t know/Depends”, “Unimportant”, and “Should not be included”. Statements rated by ≥80% as “Essential” or “Important” were designated as having consensus and were not subject to further rating. Statements rated by <70% as “Essential” or “Important” were discarded. Statements rated by 70-79% as “Essential” or “Important” were re-rated in a subsequent round with anonymised feedback. The statements were presented within 12 core constructs for ease of comprehension: (i) Basics of the mammalian circadian system; (ii) Circadian health and disruption; (iii) Assessing circadian rhythms in clinical practice; (iv) The chronobiology of BD; (v) Chronotherapy: General concepts (vi); Sleep and circadian hygiene; (vii) Bright light therapy; (viii) Wake therapy; (ix) Dark therapy; (x) Interpersonal and Social Rhythm Therapy; (xi) Melatonin and melatonergic agonists; and (xii) Cognitive Behaviour Therapy for Insomnia adapted for BD.

Round 2 included statements from Round 1 needing re-rating or rephrasing for clarity/accuracy, and new statements suggested in Round 1. A summary of the ratings for the ‘borderline’ statements from Round 1 (i.e., those rated as essential or important by 70-79% of the experts) was presented, and experts could choose to retain their original rating or change it. Round 3 included statements from Round 2 requiring re-rating; no new statements were permitted in Round 2.

### Review for acceptability

Some technical language used in clinical practice related to BD has been described as stigmatising. We invited two young persons with lived experience of BD to review the statements (∼50% each) and provide feedback, with attention to bias, language, and appropriateness.

### Statistical analysis

Analyses were conducted using R (version 4.3.3).

## RESULTS

### Expert consensus panel

The characteristics of the 30 experts who completed the study (100% of whom were retained from Round 1) are reported in the appendix (p. 3). The mean age was 55.3-years (SD=11.8); 40% were female. The sample was enriched with strong expertise, including ∼26 years (SD=10.8) of clinical experience and ∼22 years (SD=12.6) of research experience; most experts were psychiatrists (83%). Most identified as White (87%), but there was representation from 15 countries in Asia, Europe, North and South America, and Oceania.

#### Expert consensus statements

The experts reached a consensus (≥80% agreement) on 342 statements that are essential for clinicians supporting people with BD to know about the chronobiology and chronotherapeutics of BD. These consensus statements spanned four key themes (discussed below): (1) basic circadian science; (2) circadian health and disruption; (3) chronobiology of BD; and (4) chronotherapies.

Figure 1 illustrates the flow of statements across the study. Figure S1 gives a summary of the distribution of ratings across all statements examined (appendix p. 4). Figure 2 shows the distribution of the statements across the 12 core constructs.

**Figure 1.**
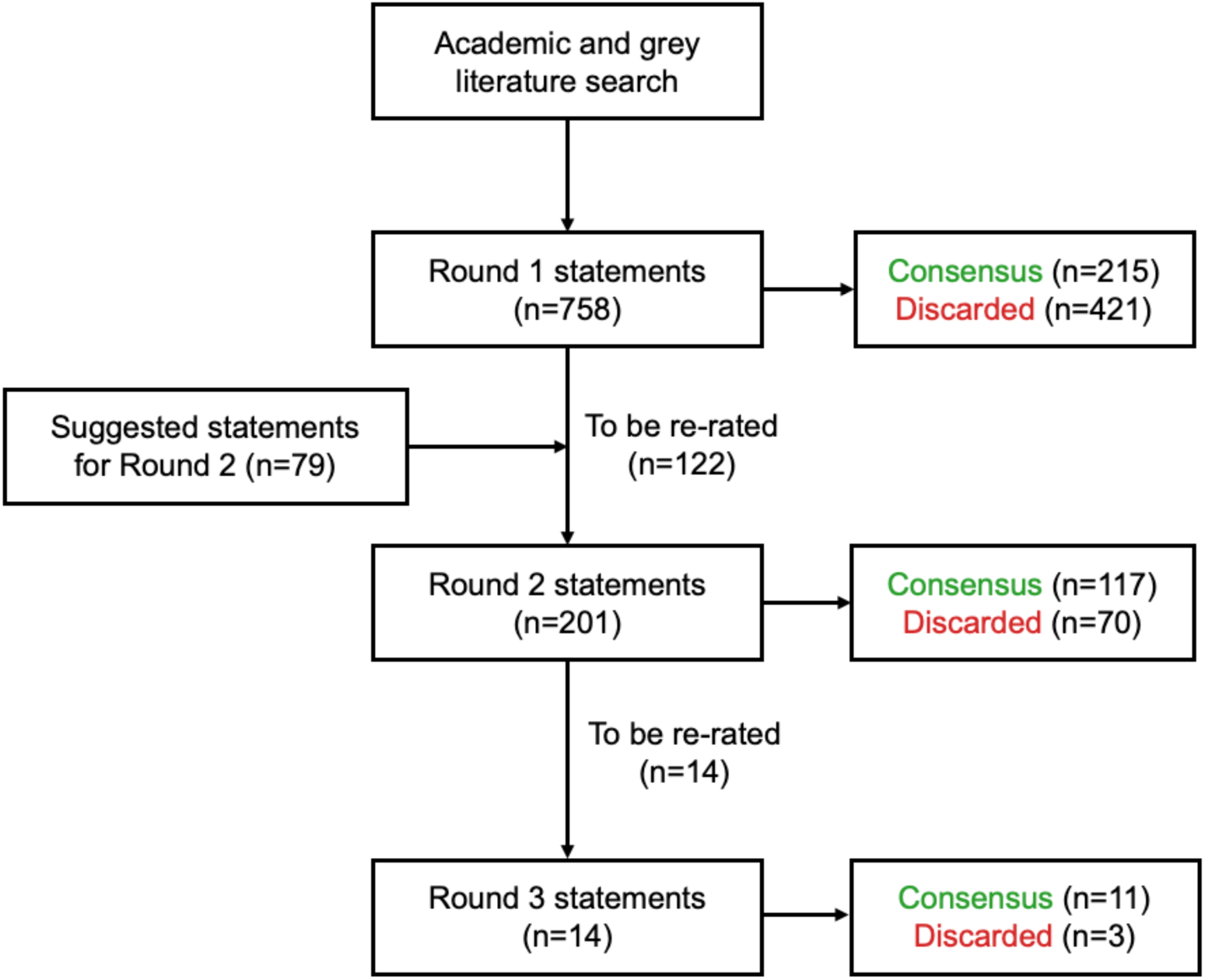
Flow of statements across the Delphi study (n=30 experts). A total of 342 statements reached consensus after all three rounds.

**Figure 2.**
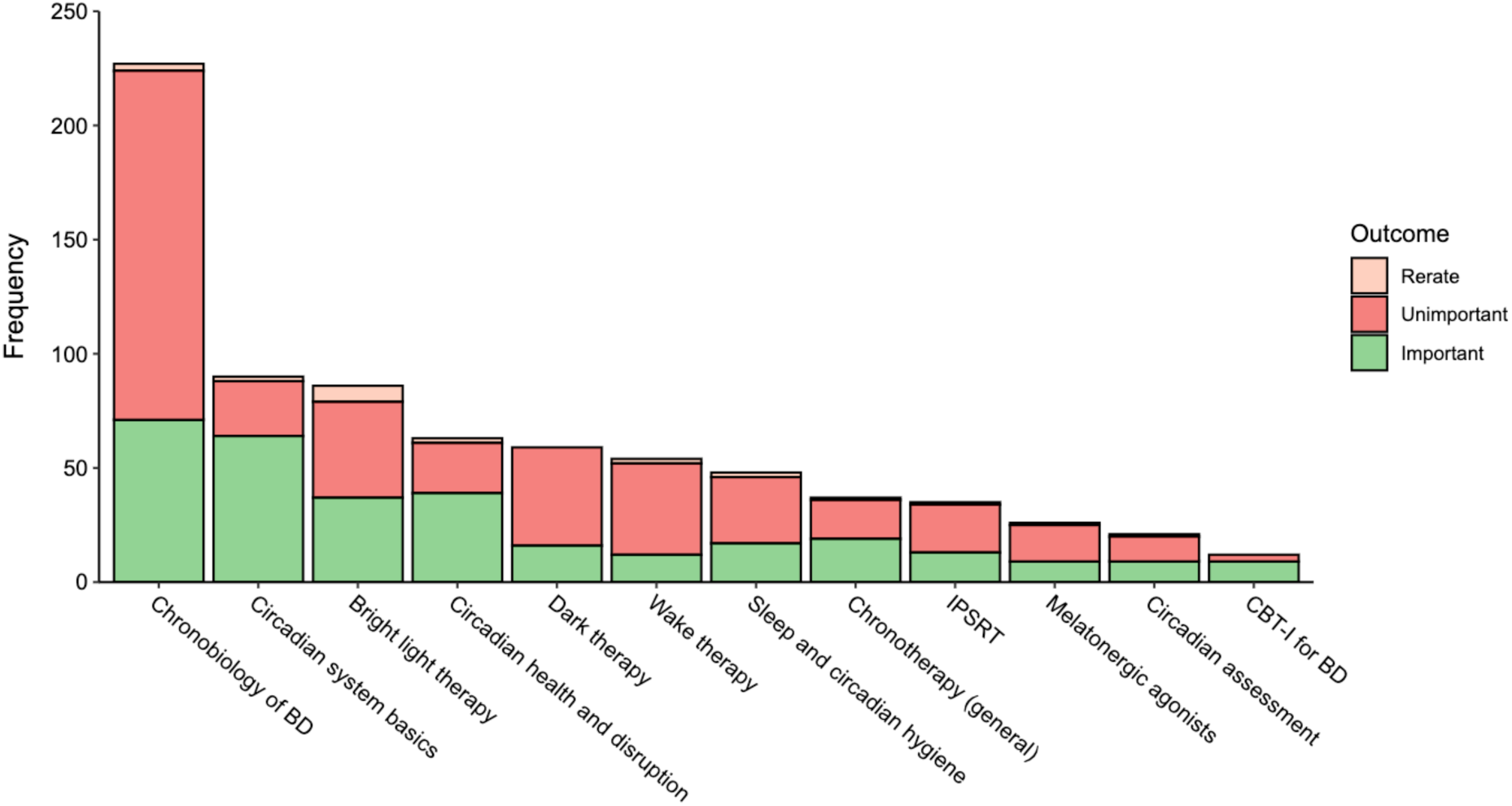
Patterns of ratings of statements categorised within core constructs. Note: The 12 constructs were created by the investigators to ease cognitive load when rating, and the columns are ordered according to the number of statements within each construct.

Given the constraints on space, we detail in Table 1 the top 60 consensus statements. These 60 represent one-sixth (17.5%) of the total 342 consensus statements; we encourage readers to examine the total set, available online (https://osf.io/agk9y). We provide links to access the consensus statements organised according to the 12 core constructs (Table 2).

**Table 1.** Summary of 60 consensus statements, selected to have representation of five statements from each of the 12 core constructs. Note:% indicates the proportion of experts rating statement as Essential/Important.

| **Core construct 1: Basics of the mammalian circadian system** | **%** |
| --- | --- |
| The suprachiasmatic nucleus (SCN) has an intrinsic rhythm which is constantly entrained by external stimuli | 100 |
| Some people's sleep-wake rhythm starts later (e.g., night owls) than others (e.g., early-birds or morning larks) | 100 |
| The light/dark cycle is the strongest zeitgeber in humans | 100 |
| Melatonin synthesis is inhibited by exposure to light | 100 |
| Melatonin is a chemical signal of darkness | 97 |
| **Core construct 2: Circadian health and disruption** |  |
| Increasing morning light exposure can shift the biological clock to an earlier phase and lessen social jetlag | 100 |
| Decreasing evening light exposure can shift the biological clock to an earlier phase and lessen social jetlag | 100 |
| Irregularity of the sleep/wake cycle is associated with greater mood instability | 100 |
| Circadian rhythm disorders can be caused by internal factors (e.g., genetic variation) or external factors (e.g., shift work, jet lag) | 97 |
| The relationships between the sleep-wake cycle and mental health are bi-directional | 93 |
| **Core construct 3: Assessing circadian rhythms in clinical practice** |  |
| Discussion about chronotype should be a regular part of psychoeducation, and can introduce people with bipolar disorder to the area of circadian rhythms and its relevance to mood disorders | 90 |
| Melatonin rhythms are one of the most direct and robust indicators of suprachiasmatic nucleus (SCN) timing | 87 |
| Actigraphy can assess the timing and amplitude of the 24-hour rest-activity rhythm | 87 |
| When there are significant differences between weekday and weekend rest-activity behaviour, this is called 'social jetlag' | 87 |
| Some output rhythms are direct or pure expressions of suprachiasmatic nucleus (SCN) activity (e.g., melatonin secretion, core body temperature, cortisol), while others are products of circadian and social-behavioural processes (e.g., rest-activity rhythm) | 87 |
| **Core construct 4: Chronobiology of BD** |  |
| Symptoms of BD can interfere with the ability to sleep | 100 |
| Seasonal changes, like changes in day length, are a common trigger for mood changes and mood episodes in BD | 100 |
| Situations involving rapid changes in timing of the ambient light/dark cycle (e.g., international travel, shift work, stimulants, childbirth) can trigger mood symptoms and episodes in BD | 100 |
| A malfunctioning body clock may underlie the mood cycles in Bipolar Disorder | 97 |
| Poor sleep can worsen symptoms of BD | 97 |
| **Core construct 5: Chronotherapy: General concepts** |  |
| Chronotherapies are techniques that affect circadian rhythms and the central circadian clock | 97 |
| Chronotherapies for affective disorders are treatments that seek to alter mood-states by affective circadian rhythms and the central circadian clock |  |
| Chronotherapies use manipulation of sleep, wake, and light | 97 |
| The 5 major modalities of affective chronotherapies are: bright light therapy; dark therapy; melatonin and melatonergic agonists; treatments using sleep deprivation or wake therapy; interpersonal and social rhythm therapy; and cognitive behavioural therapy for insomnia adapted for BD | 97 |
| The integrated treatment approach of BD should include attention to the 24 hour light/dark cycle as well as focus on the sleep-wake cycle | 97 |
| **Core construct 6: Sleep and circadian hygiene** |  |
| People with BD should get out of bed at the same time each day | 97 |
| To sleep well, people with BD should avoid stimulants (e.g., caffeine, nicotine, chocolate) late in the day | 97 |
| For people with BD, it may be helpful to change their attitude to sleep (e.g., welcome it as a contributor to wellbeing and not wasted time) | 93 |
| People with BD should turn off screens, phones, and other devices 1-2 hours before bedtime | 93 |
| Regular routines and sleep schedules may lengthen the time between manic or hypomanic episodes | 93 |
| **Core construct 7: Bright light therapy** |  |
| Timing of exposure affects the outcome and tolerability of bright light therapy | 97 |
| Distance from the light device affects the outcome and tolerability of bright light therapy | 97 |
| When using bright light therapy, provided that a person with BD is treated with appropriate mood-stabilising medications and has no additional risk factors, the risk of switching into hypomania or mania appears to be relatively rare | 97 |
| Bright light therapy has strong evidence as an antidepressant for bipolar depression | 93 |
| Bright light therapy is well-tolerated in BD | 93 |
| **Core construct 8: Wake therapy** |  |
| There is a slight risk of switching into mania for a person with BD receiving wake therapy | 90 |
| Sleep deprivation therapies work rapidly | 87 |
| By themselves, sleep deprivation therapies lead to only transient improvements in mood | 87 |
| Therapeutic sleep deprivation is typically combined with medication or other chronotherapies, particularly bright light therapy and sleep phase advance | 87 |
| When used for acute bipolar depression, therapeutic sleep deprivation should always be combined with mood stabilising medication | 83 |
| **Core construct 9: Dark therapy** |  |
| Modern dark therapy involves wearing special orange tinted glasses which block the blue wavelength from artificial and natural light | 100 |
| “Virtual darkness” can be created by wearing blue light blocking glasses | 97 |
| When not in a manic episode, blue-blocking glasses can be worn at a regular time, about 2 hours before bed, in order to promote regular endogenous melatonin-production and better sleep | 93 |
| Blue-blocking glasses may be useful for treating mania in BD | 90 |
| A person with BD can darken their bedroom using black-out curtains | 90 |
| **Core construct 10: Interpersonal and Social Rhythms Therapy** |  |
| In Interpersonal and Social Rhythm Therapy (IPSRT), people learn to identify and manage events that destabilise their daily routines | 100 |
| In IPSRT, people learn to track the regularity of 5 activities: (1) time out of bed; (2) first contact with another person; (3) time starting formal commitments; (4) dinner; and (5) bed-time | 97 |
| In IPSRT, the Social Rhythm Metric is used to track and improve the regularity of social activities | 90 |
| IPSRT improves functioning in people with BD | 90 |
| IPSRT works for BD because people with BD are vulnerable to dysregulation of daily rhythms | 83 |
| **Core construct 11: Melatonin and melatonergic agonists** |  |
| There is insufficient evidence to endorse or reject melatonin for the acute phase of bipolar depression | 87 |
| There is insufficient evidence to endorse or reject melatonergic agonists for the acute phase of bipolar depression | 87 |
| There is insufficient evidence to endorse or reject melatonergic agonists for prophylaxis of bipolar depression | 83 |
| Taking melatonin an hour or so before the desired bedtime sends an entraining signal to the circadian clock | 83 |
| Melatonin quality is variable, suggesting the need for use only of approved products | 83 |
| **Core construct 12: Cognitive behaviour therapy for insomnia adapted for BD** |  |
| Cognitive behaviour therapy for insomnia (CBT-I) addresses thoughts and beliefs that interfere with sleep and teaches sleep hygiene practices | 97 |
| CBT-I for BD (CBT-I-BD) aims to improve mood, sleep, and functioning in people with BD | 97 |
| CBT-I may improve insomnia in BD and other mood disorders | 90 |
| CBT-I-BD produces benefits for sleep after acute treatment among people with inter-episode BD and comorbid insomnia | 87 |
| CBT-I-BD produces benefits for mood episode relapse after acute treatment among people with inter-episode BD and comorbid insomnia | 87 |

**Table 2.** Consensus statements organised according to 12 core constructs.

| Core constructs | Access the statements |
| --- | --- |
| 1. Basics of the mammalian circadian system | https://osf.io/ktq2x |
| 2. Circadian health and disruption | https://osf.io/pnuxg |
| 3. Assessing circadian rhythms in clinical practice | https://osf.io/qg3pc |
| 4. The chronobiology of bipolar disorder | https://osf.io/d4rmq |
| 5. Chronotherapy: General concepts | https://osf.io/pgj4w |
| 6. Sleep and circadian hygiene | https://osf.io/f3acd |
| 7. Bright light therapy | https://osf.io/mz7kd |
| 8. Wake therapy | https://osf.io/kc7nx |
| 9. Dark therapy | https://osf.io/g4qjv |
| 10. Interpersonal and Social Rhythms Therapy | https://osf.io/mx8ub |
| 11. Melatonin and melatonergic agonists | https://osf.io/ctfkx |
| 12. CBT for insomnia adapted for Bipolar disorder | https://osf.io/6eqwp |

#### Consensus Theme 1: Basic circadian science

It is essential that clinicians know about basic aspects of circadian science. Consensus was reached on 70 statements in this theme. All experts agreed that clinicians should know that the pacemaker of the circadian system (the suprachiasmatic nucleus [SCN]) has an intrinsic rhythm that is constantly entrained by external stimuli (e.g., light); that some people’s sleep-wake cycle starts later (night owls) than others (morning larks); and that melatonin synthesis is inhibited by exposure to light (all but one expert thought that clinicians should know that melatonin is the ‘chemical signal of darkness’; this is in contrast to the popular idea that melatonin is a sleep hormone).

Other consensus statements included topics spanning definitions (e.g., phase, period) and types of circadian rhythms (e.g., core body temperature, cortisol); that circadian rhythms are widespread throughout the body and are found in almost all cells; that the circadian system regulates many processes relevant to BD (e.g., mood, cognition, hormone secretion, gene expression); that circadian rhythms can shift earlier and later relative to clock time; that zeitgebers (German for “time-givers”) entrain the circadian system (with zeitgebers being the timing of light, eating, work, exercise, social activity); that melatonin is a key circadian; that there are direct effects of light on brain areas relevant to mood and cognition; that the circadian system is most sensitive to short-wavelength “blue” light (460-480nm); and that “blue” light promotes wakefulness and suppresses melatonin.

Consensus in this theme may reflect the idea that if clinicians are aware of the ‘what’, ‘how’, and ‘why’ of the circadian system, then they may be more motivated to engage with circadian principles and to correctly employ the chronotherapies in Theme 4.

#### Consensus Theme 2: Circadian health and disruption

It is essential that clinicians know about the causes of circadian disruption and consequences for health and wellbeing. Consensus was reached on 42 statements related to this theme. All experts agreed that clinicians should know that increasing morning light exposure and decreasing evening light exposure can shift the timing of the circadian clock earlier, and that irregularity of the sleep-wake cycle is associated with greater mood instability. Other consensus statements were that circadian rhythm disorders are multifactorial and can be caused by internal (e.g., genetic variation) and external factors (e.g., shift work, jet lag); that the relationships between the sleep-wake cycle and mental health are bi-directional; that circadian disruption is associated with risk of mood instability and mood disorders; that evening chronotype is associated with higher risk of mood disorders; and that knowing one’s chronotype can help someone understand their internal clock and how to synchronise it.

Consensus within this theme may reflect two ideas: (1) clinicians should know about circadian disruptors and how people with BD can minimise disruption by changing their behaviour and environments; and (2) clinicians should be equipped with principles that can help them predict the illness course of their patients (e.g., seasonal patterns).

#### Consensus Theme 3: Chronobiology of BD

It is essential that clinicians know about key aspects of the chronobiology of BD. Consensus was obtained on 73 statements about the chronobiology of BD and 11 statements about assessment of circadian rhythms in clinical practice. Consensus statements included the role of circadian processes in the etiology and course of BD; that the phenomenology of BD encompasses processes regulated by circadian rhythms (e.g., mood, appetite, energy); that sleep disturbance is a prognostic signal in the prodrome of mania; that sleep disturbance often endures inter-episodically in BD; that people with BD may have altered sensitivity to zeitgebers (time cues) like light; that there are bidirectional relationships among symptoms of BD and sleep, the sleep-wake cycle, and circadian rhythms; that factors that disrupt the sleep-wake cycle (e.g., international travel, shift work, childbirth) affect mood changes in BD; that there is meaningful inter-individual variability in sleep and circadian features in BD; that sleep disruption is associated with serious clinical outcomes (e.g., suicidality); that people with BD may be hyper-sensitive to the circadian-disrupting effects of artificial light at night; that stabilisation of circadian rhythms is a therapeutic mechanism in BD; that lithium can phase shift circadian rhythms; and that stabilisation of circadian rhythms may be an essential mechanism of certain therapies (e.g., lithium).

Consensus within this theme may reflect the notion that the circadian system is implicated in many aspects of BD and that improving circadian rhythmicity is a logical therapeutic strategy.

#### Consensus Theme 4: Chronotherapies

Finally, it is essential that clinicians know about the six major affective chronotherapies, including the evidence in support of them and information about procedures (e.g., dose, timing), outcomes, side effects, and contraindications. Comments from the experts on specific statements emphasised a need for more rigorous clinical trials, and post-study correspondence highlighted that this consensus statement would lay the foundation for future collaborative work in clinical trials.

##### Bright light therapy (BLT)

This chronotherapy had the most consensus statements (n=60). In addition to the statements in Table 2, clinicians should know about key parameters that affect the outcome and tolerability of BLT (e.g., intensity of light [lux], wavelength, device angle, exposure duration); BLT should provide 7,000-10,000 lux at a distance of 30-38cm (12-15 inches), for 30-120 min per day (depending on light output); that for the depressed phase of BD, midday administration may be effective, while nighttime administration is not recommended; adverse effects appear to be mild; and that a trained clinician can recommend which type of device can be used, and that BLT should be used under clinical supervision.

##### Wake therapy

This chronotherapy had 12 consensus statements. In addition to those in Table 2, clinicians should know that wake therapy should only be done under clinical supervision; that adding BLT to wake therapy can prolong the therapeutic effect; that wake therapy is used only during the depressed phase of BD because it can worsen or trigger manic symptoms; and that wake therapy may not be suitable for people with comorbid epilepsy (it may induce seizures).

##### Dark therapy

Most of 20 consensus statements were related to modern dark therapy (blue-blocking glasses [BBGs]). Clinicians should know that dark therapy is not a substitute for medication in BD; that BBGs should block 90% of blue light; that BBGs can improve sleep efficiency and lead to a rapid resolution of mania in hospitalised people; that while manic, people with BD should wear BBGs from ∼6pm until getting into bed and turning off lights, and again in the morning if they are awake, until 8am; and that people with BD can reduce their environmental exposure to light at night using an eye mask or black-out curtains.

##### Interpersonal and Social Rhythms Therapy (IPSRT)

There were 13 consensus statements about IPSRT. In addition to statements in Table 2, clinicians should know that IPSRT and CBT can be combined together and used with other mood stabilizers and antipsychotics; that IPSRT appears feasible to deliver in people with BD and to youth at-risk of BD; that IPSRT is a promising adjunctive for adolescents with BD, and Social Rhythm Therapy can improve mood in youth with BD; and that IPSRT reduces depressive symptoms in people with BD.

##### Melatonin and melatonergic agonists

Most of the 10 consensus statements indicated that that there is insufficient evidence to endorse or reject melatonin or melatonergic agonists for the acute phases or prophylaxis of bipolar depression and mania in BD.

##### Cognitive behaviour therapy for insomnia adapted for BD (CBT-I-BD)

There were 9 consensus statements related to this chronotherapy. Clinicians should know about the structure and aims of CBT-I and CBT-I-BD, and that CBT-I-BD can improve sleep and reduce relapse after acute treatment in people with inter-episode BD and comorbid insomnia, and that a package that combines CBT-I, psychoeducation, and light-dark cycle management can improve sleep and mental health in inpatients with BD.

## DISCUSSION

This study identified a core knowledge base of chronobiology and chronotherapeutics for clinicians, which is intended to support and optimise their care of people with BD. Using a Delphi process with 30 experts from 15 countries, ∼750 statements (collated from an exhaustive search of scientific and grey literatures) was distilled to a final set of 342 statements. This process unpacks the chronobiologic model and its application to BD into a series of practices and principles that will enable this paradigm to be more effectively employed in the understanding and treatment of BD.

### Dissemination

Dissemination and implementation science studies how information is distributed and assimilated by clinicians. It seeks to determine the ‘how, what, when, where, and to whom’ findings are best incorporated and utilised in clinical practice. To determine the optimal strategies for the translation of this Delphi-based chronobiologic and chronotherapeutic information, we must consider several factors including the content and size of the study’s final set. Three-hundred-and-forty-two statements were rated as critical by our experts, forming a large educational package. We did not impose an *a priori* limit on its scope, allowing experts to choose and rank statements without concern for the ultimate size of the set. Given its large size, how can this information be optimally organised, and through what channels can it be distributed to most effectively educate clinicians?

First, we (along with the experts; Figure S2 in Appendix) believe that the clinical uptake and utilisation of this chronobiologic and chronotherapeutic model is best supported through its incorporation into CPGs and training curricula. As one of the now-foundational models of the pathophysiology of BD, meaningful dissemination of the science requires inclusion of this area in graduate and postgraduate education and CPGs. The breakdown of these consensus statements into four major themes (basic circadian science; circadian health and disruption; chronobiology of BD; and chronotherapies) lends itself to a several-session educational series, whether through a short course of classes, webinars, or Continuing Medical Education materials.

This information could also be further refined and tailored to specific settings. For example, clinicians with different training may benefit from learning different levels of this knowledge base. While psychiatrists might ideally employ the full breadth of these principles and practices, psychologists, nurses, and primary care physicians may require a condensed version (see Table S2 in Appendix for a heuristic framework for how information might be tailored to clinician types). The educational program could be customized for specific clinical situations such as the initial assessment and formulation, management of acute episodes, maintenance therapy, and treatment and prevention of specific timing misalignments (e.g. seasonal change, shift work).

Another question bearing on effective dissemination is whether this information is best organised as a stand-alone circadian model of care or whether these practices and principles would be better included in existing, evidence-based therapies such as psychoeducation or IPSRT. Recent proposals of circadian types of depression would support the employment of pathophysiology-specific, stand-alone interventions^19^. In contrast, we believe that many of the consensus-endorsed statements obtained here are so important and widely relevant that they should be part of the standard of care for all clinicians working in mood disorders (Table S2, Appendix). This would include, for example, the need to manage the timing and degree of light and dark exposure, or the relevance of a patient’s chronotype to understanding sleep disturbances and in tailoring chronotherapeutic interventions. Other items, such as how to implement the less common chronotherapies (e.g., wake therapy), might be reserved for specialised classes or educational forums.

There have been major innovations in the past few years in the estimation of circadian parameters in real-world settings via application of mathematical models to wearables^41,42^. The combination of improved knowledge among clinicians with real-time continuous measurement of behaviour^22,43,44^, and models that can estimate circadian rhythms and deliver this information digitally may offer new opportunities to personalise and scale chronotherapeutic management strategies for people with BD.

### Limitations

First, our recruitment strategy failed to attract strong representation of three kinds of experts: (a) residents of the African continent and two of the most populated countries (India, China); (b) residents of several countries facing circadian challenges caused by their latitude (e.g., Finland, Iceland); and (c) non-psychiatrist health professionals that play a role in the management of BD (e.g., psychologists, general practitioners, mental health nurses). This may bias the generalisability of the consensus information and may have prevented rating of information specific to these contexts. Second, given the diversity and complexity of this area, many statements were rated in Round 1. This was burdensome and, in some cases, caused attrition. Third, given that most experts are members of a task force dedicated to the chronobiology and chronotherapy of BD, it is possible that the consensus information is biased toward greater inclusiveness of information, as compared to what might have been derived by generalists or specialists in other areas of BD. Finally, only two people with lived experience of BD reviewed the statements in Round 1. Including more people with lived experience and thereby capturing conflicting experiential knowledge may have uncovered novel insights about the value and limitations of this chronobiological model of BD.

## Conclusion

In conclusion, we obtained an expert consensus about concepts, principles, and practices related to the chronobiology and chronotherapy of BD, which experts think are essential for clinicians to know to optimise their care of people with BD. Our next step will be to organise and disseminate this information, similar to the Society for Biological Rhythms “Circadian Medicine Course”^35^ but with a BD focus. We expect that the uptake of this consensus information will improve the outcomes of people with BD, especially if the findings are incorporated in CPGs and clinical training programs^24^. We also invite readers to use the full consensus body of information (https://osf.io/agk9y) in their own educational programs.

## Data Availability

The data are available upon request to Dr Jacob Crouse

## Contributor Roles Taxonomy (CRediT)

**Conceptualisation:** JJC, JFG

**Data curation:** JJC, VL, TW, and JFG

**Formal analysis:** JJC, VL, TW, and JFG

**Funding acquisition:** JJC

**Investigation**: JJC, VL, JFG

**Methodology:** JJC, VL, AJ, and JFG

**Project administration:** JJC, VL, and JFG

**Software:** JJC and TW

**Visualization:** JJC and TW

**Writing – original draft:** JJC and JFG

**Writing – review & editing:** All authors

